# Expert-Guided Visual Correction for Characterizing Diagnostic Performance and Error Patterns of Multimodal Large Language Models Using Periodontal In-Service Examination Images

**DOI:** 10.64898/2026.08.21.26360755

**Authors:** Prita Abhay Dhaimade, Robin Henderson

## Abstract

Multimodal large language models (MLLMs) are increasingly applied to image-based clinical reasoning, yet their diagnostic reliability in periodontal image interpretation, and the underlying source of their errors, remain poorly characterized. This study evaluated six architecturally distinct MLLMs (Claude Sonnet 4.5, GPT-5.0, Gemini 2.5, GLM-4.6, Sonar, and Grok 4.1) using 50 image-based multiple-choice questions drawn from the American Academy of Periodontology In-Service Examination, spanning clinical photographs, histopathology, radiographs, cardiac rhythm strips, and anatomical illustrations. A sequential two-phase experimental design was used: in Phase 1, each model independently described each image, selected an answer, and provided a supporting citation; in Phase 2, applied only to questions answered incorrectly, models were given an expert-validated visual description and asked to re-answer, allowing diagnostic improvement through visual correction to be measured directly. Expert ground truth for image content was established by a board-certified periodontist and independently validated by a second board-certified periodontist. Model outputs were classified using a dual-process error taxonomy adapted from Norman’s model of diagnostic reasoning, distinguishing perceptual errors, arising from inaccurate visual feature extraction, from cognitive errors, arising from flawed reasoning despite accurate perception, with cognitive errors further subdivided into correctable and persistent subtypes, and additional categories capturing compound perceptual-cognitive failures and compensatory reasoning that overcame inaccurate perception. Diagnostic accuracy and error type distribution varied significantly across models and image modality. Correcting inaccurate visual descriptions in Phase 2 improved diagnostic accuracy for a subset of previously incorrect responses, indicating that a meaningful share of errors originated at the level of visual perception rather than clinical reasoning; conversely, a distinct subset of errors persisted despite accurate corrected visual input, indicating reasoning-level failures independent of perceptual accuracy. Some models also reached correct answers despite generating inaccurate image descriptions, reflecting compensatory reasoning resilient to perceptual error. These findings show that aggregate accuracy scores conflate mechanistically distinct failure modes, and that perceptual and cognitive errors carry different implications for how MLLMs might be safely deployed or improved for diagnostic image interpretation. The expert-guided visual correction framework introduced here provides a generalizable, mechanism-based approach to benchmarking multimodal AI diagnostic performance that extends beyond periodontics to other visually driven diagnostic domains in medicine. As MLLMs become increasingly accessible to clinicians, residents, and dental educators, distinguishing perceptual from cognitive failure is essential for guiding responsible clinical use, targeting model refinement, and informing AI-augmented dental education and competency assessment.

## Introduction

Multimodal large language models (LLMs) integrating vision and language capabilities have demonstrated promising performance in medical image interpretation across radiology, pathology, and dermatology. (1–3) These systems process complex visual information and generate diagnostic assessments through sophisticated vision encoders, multimodal fusion mechanisms, and transformer-based reasoning architectures. (2,4–7)

Central to this pipeline is a learnable multimodal connector that bridges vision and language components. (1) Modality-specific encoders convert complex data forms like images, audio, and video into smaller and manageable informative representations by identifying and extracting key features. The mapping of visual encoder outputs into language-compatible representations varies across architectures based on connector type and its fidelity in preserving visual information. (1,7)

As clinical deployment of these systems becomes increasingly feasible, understanding not only whether they succeed or fail, but *why* and *where* in their processing pipeline failures occur, is critical for safe clinical implementation. (6,8) Yet systematic evaluation of the mechanistic origins of diagnostic errors, whether arising from perceptual limitations in visual feature extraction or cognitive deficits in reasoning, despite accurate perception, remains largely unexplored in contemporary multimodal AI benchmarking studies.

Recent studies have systematically benchmarked visual and multimodal LLM diagnostic accuracy across diverse medical imaging tasks, including chest radiography, board-style surgical imaging questions, radiolucent jaw lesions, and NEJM (New England Journal of Medicine) Image Challenge cases. The primary focus of a number of these studies remains aggregate performance metrics, without distinguishing the locus of failure. (9–12) While studies have employed example-guided prompting with reference images to improve oral lichen planus detection or classified multimodal LLM errors observationally across image comprehension, (13) systematic evaluation of whether providing expert-corrected visual descriptions for the same image improves diagnostic accuracy remains limited. Such an intervention would be critical for isolating perceptual failures from persistent reasoning deficits. This distinction has important implications for model development: perceptual errors may respond to improved vision encoders or medical imaging-specific pre-training, whereas cognitive errors suggest limitations in diagnostic reasoning frameworks that may require different optimization strategies. Looking at the available research material, recently numerous studies have benchmarked large language model (LLM) accuracy in dental education and periodontics, yielding performance ranges of 60-80% on standardized assessments including the American Academy of Periodontology (AAP) In-Service examination and structured clinical periodontal queries. (14–17)

These investigations, however, have been confined to static baseline evaluations of closed-ended question formats, without probing intervention-mediated accuracy improvements or systematically appraising the fidelity of open-ended, LLM-generated interpretive content for dental imagery.

Building upon these foundational benchmarks, this study presents a systematic evaluation of perceptual versus cognitive error mechanisms in multimodal LLMs through expert-guided visual correction in periodontal image interpretation. We benchmarked six architecturally distinct multimodal LLMs across 50 image-based questions selected from the American Academy of Periodontology AAP, In-Service examination. The questions spanned across five imaging modalities (Clinical photographs, Histopathology, Radiographs, Cardiac rhythm strips and Anatomical illustrations). The protocol used consisted of two distinct phases: initial diagnostic assessment followed by conditional intervention providing expert visual descriptions when errors occurred. This design enabled empirical distinction between correctable perceptual errors and persistent cognitive failures, characterization of domain-specific failure patterns, and identification of architectural features associated with correction responsiveness addressing critical gaps in prior work that examined only static baseline accuracy without intervention effects or open-ended content evaluation.

## METHODS

### 1. Study Design

This cross-sectional benchmarking evaluation employed a sequential two-phase factorial experimental design to assess multimodal large language model (LLM) performance in periodontal image interpretation. (Fig 1) The study protocol incorporated a conditional corrective intervention component to evaluate real-time diagnostic improvement through expert-guided visual correction.

**Figure 1:**
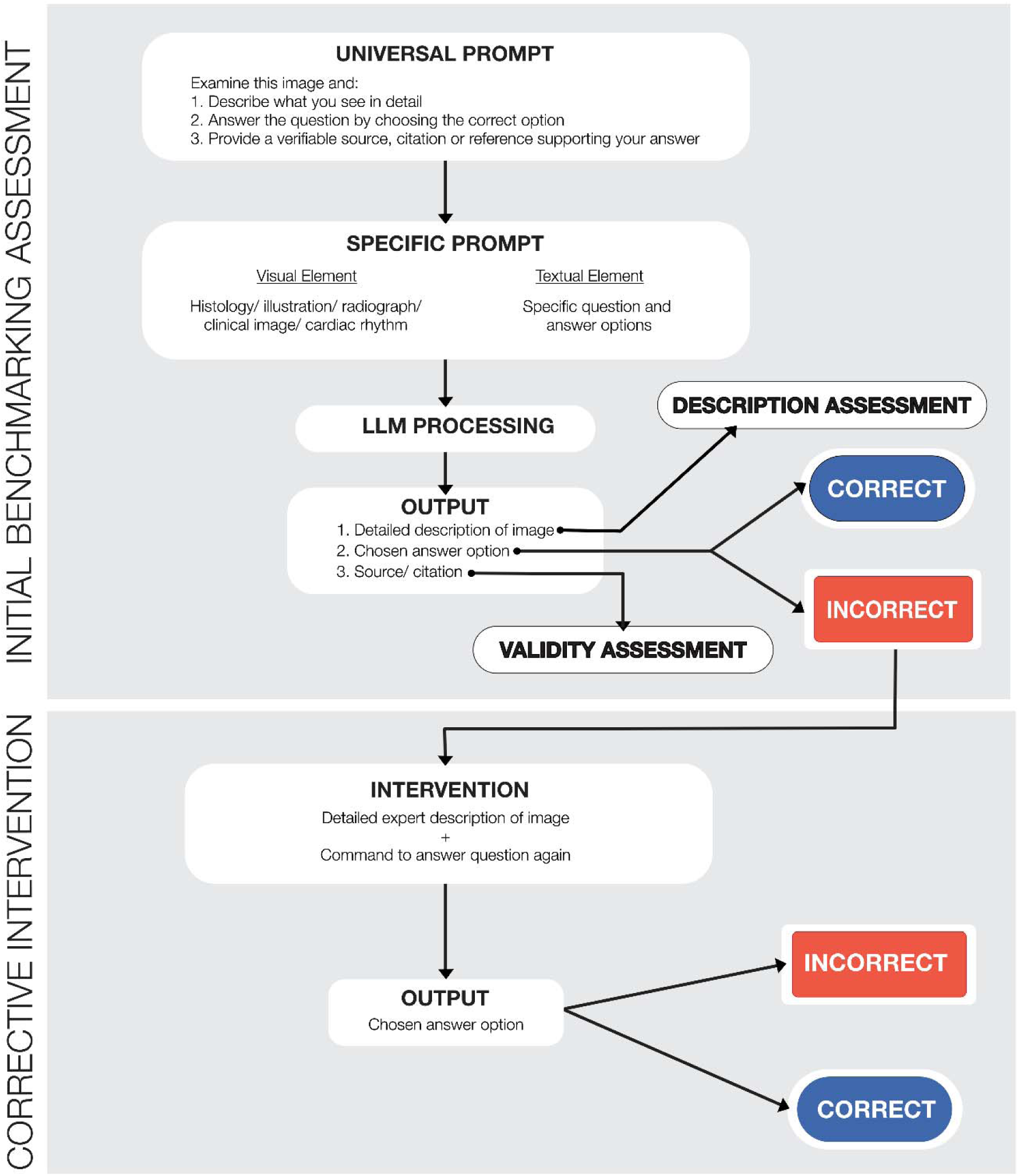
Schematic diagram of the two-phase experimental protocol. Phase 1 (Initial Benchmarking Assessment): Universal prompt with visual and textual elements processed by LLM, generating image description, answer selection, and citation. Outputs assessed for description accuracy and answer validity. Phase 2 (Corrective Intervention Assessment): Triggered when Phase 1 produces incorrect description or answer. Expert visual description provided with prompt to re-answer question. Output assessed for correctness.

### 2. Evaluation Dataset

#### American Academy of Periodontology In-Service Examination as Educational Instrument

The American Academy of Periodontology (AAP) In-Service Examination is administered annually to periodontal residents across all three years of specialty training in Commission on Dental Accreditation (CODA)-accredited periodontics programs throughout the United States.

The examination serves as a formative assessment tool designed to evaluate progressive knowledge acquisition and benchmark resident performance against national cohorts at equivalent training levels rather than as a licensure or board certification requirement. Questions span foundational biomedical sciences (embryology, anatomy, biochemistry, physiology, microbiology, immunology) through advanced Clinical photograph domains (periodontal diagnosis, treatment planning, surgical therapy, pharmacotherapeutics, oral pathology).

The AAP provides individual residents with confidential performance reports showing percentile rankings relative to same-year peers nationally. However, the AAP does not publicly release aggregate performance statistics, mean scores, or score distributions to maintain examination security and protect resident confidentiality.

### 3. Dataset Construction for Present Study

Fifty image-based multiple-choice questions were systematically selected from available AAP

In-Service Examination education materials spanning examination years 2012–2025. Questions were categorized by the study authors according to image type into five distinct modalities:

- **Clinical photographs:** 21 questions (42%) depicting soft tissue presentations, periodontal defects, gingival pathology, and clinical findings
- **Histopathology:** 10 questions (20%) showing microscopic tissue architecture, cellular patterns, and histological pathologic entities
- **Radiographs:** 7 questions (14%) including periapical, bitewing, panoramic images and other radiographs
- **Cardiac rhythm strips:** 6 questions (12%) presenting electrocardiogram tracings requiring recognition and treatment implications
- **Anatomical illustrations:** 6 questions (12%) consisting of schematic diagrams depicting anatomical structures and morphology

Images were used in their original format without modification, cropping, annotation, or digital enhancement and all questions were presented in multiple-choice format with response options as they originally appeared in the examination.

### 4. Large Language Models Selection, Access, and Configuration

Six multimodal large language models were selected to represent architecturally distinct systems with vision capabilities, maximizing diversity across vision encoder architectures, multimodal fusion mechanisms, and training methodologies. The benchmarking panel included:

1. **Claude Sonnet 4.5** (Anthropic, San Francisco, CA, USA)
2. **GPT-5.0** (OpenAI, San Francisco, CA, USA)
3. **Gemini 2.5** (Google DeepMind, Mountain View, CA, USA)
4. **GLM-4.6** (Zhipu AI, Beijing, China)
5. **Sonar** (Perplexity AI, San Francisco, CA, USA)
6. **Grok 4.1** (xAI, San Francisco, CA, USA)

All models were accessed through standard consumer-facing web-based chat interfaces using commercially available configurations rather than through research application programming interfaces (APIs) or custom deployments. This approach was intentionally chosen to emulate how dental students, residents, and clinicians typically interact with these systems in real-world educational or commercial contexts, without specialized technical infrastructure or programming expertise. LLM level differences in processing architecture can be noted in Fig 2.

**Figure 2:**
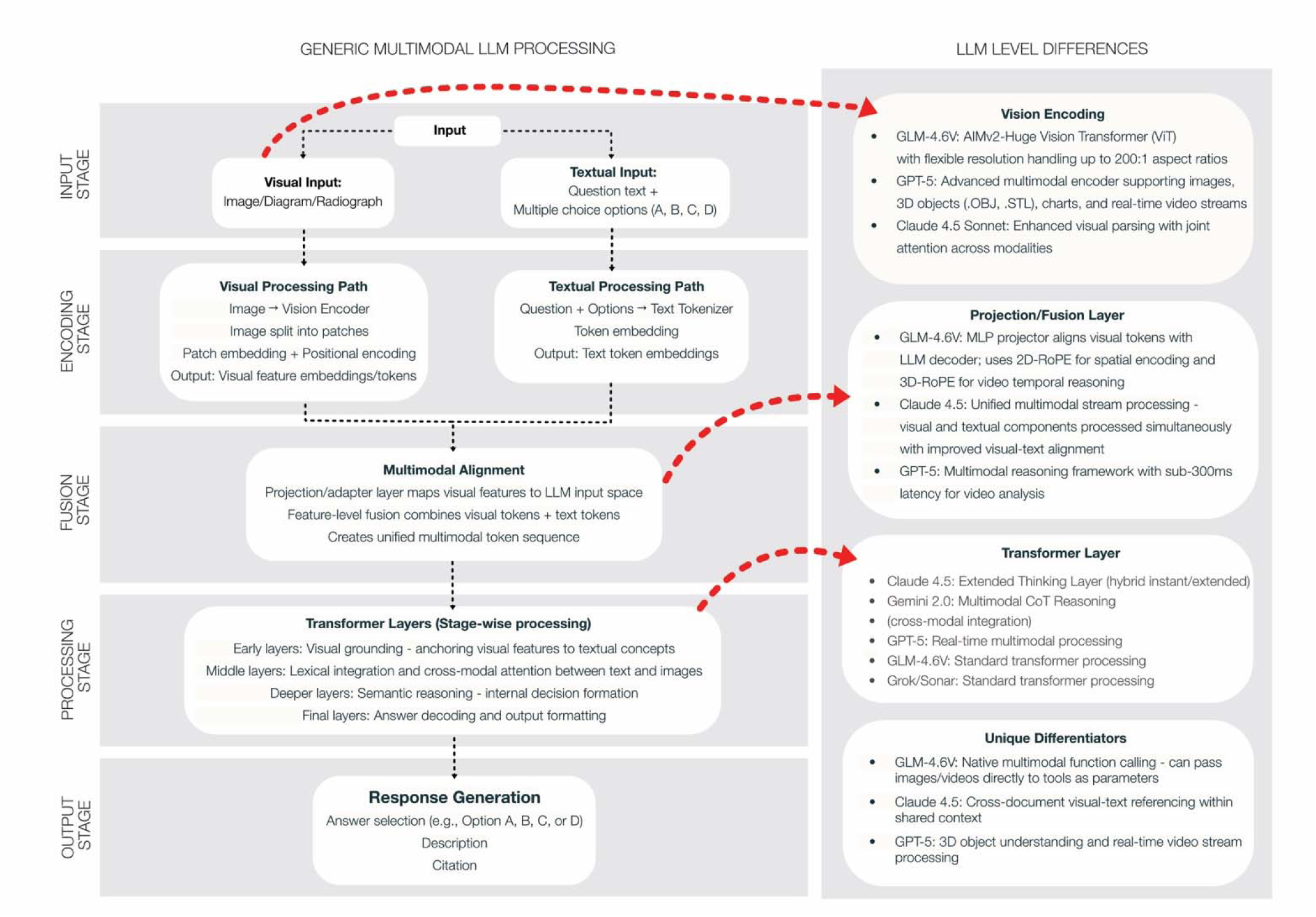
Multimodal LLM Processing Architecture for an image related question and Model-Specific Variations. Schematic of multimodal LLM processing pipeline (left) and architectural variations among evaluated models (right). Processing stages: input, encoding (parallel visual and textual paths), multimodal fusion, transformer layers, and response generation. Right panel shows model-specific differences in vision encoding, projection/fusion mechanisms, transformer implementations, and unique architectural features. Red arrows indicate key architectural variation points.

### 5. Ground Truth Establishment Answer Key Validation

Correct answers for all 50 questions were established using the official AAP In-Service Examination answer keys as published.

#### Visual Description Ground Truth

Expert visual descriptions for all 50 images were developed by a single board certified periodontist (P.D.) and independently validated by a second board-certified periodontist (R.H.), to serve dual purposes: (1) as the reference standard for evaluating Phase 1 visual description accuracy, and (2) as the corrective input for Phase 2 intervention.

For each of the 50 images, the expert evaluator documented all clinically relevant visual findings, anatomical structures, pathological features, tissue characteristics, and diagnostic signs observable in the image. These descriptors are documented in supplemental files.

### 6. Experimental Protocol

#### Phase 1: Initial Benchmarking Assessment

Each of the 50 image-based questions was presented independently to all six LLMs using an identical standardized prompt structure. To minimize cross-question contamination while preserving ecological validity, each question was entered into a new chat window or conversation thread for every model.

**Standardized Phase 1 Prompt:**

“Examine the following image and associated question.

1. *Describe in detail what you observe in the image*.
2. *Based on your visual analysis, select the single best answer to the question from the options provided*.
3. *Provide a verifiable citation or reference (with DOI/PMID/URL) that supports your answer.”*

The visual descriptions provided, answers chosen, and citations provided were documented and assessed for accuracy against expert ground truth and official AAP answer keys.

#### Phase 2: Corrective Intervention Assessment

Phase 2 was conditionally triggered to evaluate whether providing expert-validated visual descriptions could improve diagnostic accuracy through vision-language realignment.

#### Intervention Trigger

Phase 2 intervention was applied only when Phase 1 assessment for a given question gave an incorrect answer. Questions where the answer was correct in Phase 1 did not trigger Phase 2.

**Standardized Phase 2 Prompt:**

For all triggered cases, the same LLM that produced the Phase 1 response was presented with the following standardized corrective prompt in the same conversation thread:

*“Here is what should be observed in this image: [expert-validated visual description]*.

*Based on this corrected visual information, answer the question again by selecting the single best option.”*

For each triggered question–model pair, the corrected answer provided and improvement status (whether accuracy improved from Phase 1 to Phase 2) were documented.

### 7. Citation Validation Protocol

Citations provided by LLMs were collected verbatim, verified through a four-step human-in-the-loop protocol, and classified as Valid or Not Valid. Detailed validation methodology and analysis will be reported separately.

### 8. Error Classification Framework

All question–model combinations were classified using a dual-process error taxonomy based on Norman’s model, distinguishing perceptual errors (visual feature extraction failures) from cognitive errors (reasoning failures despite correct perception), with cognitive errors further categorized as correctable or persistent. (Table 1)(18,19)

**Table 1:**
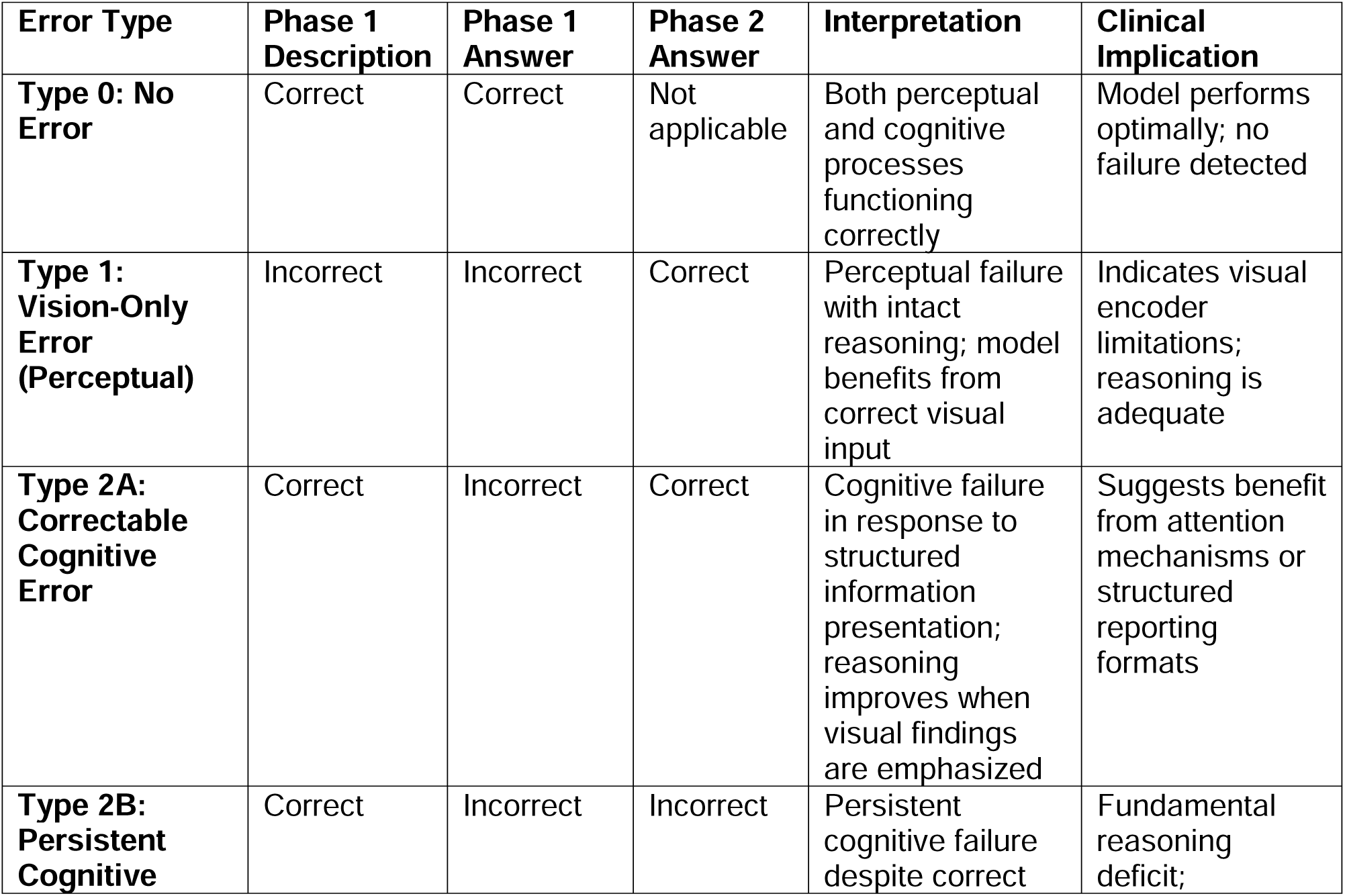
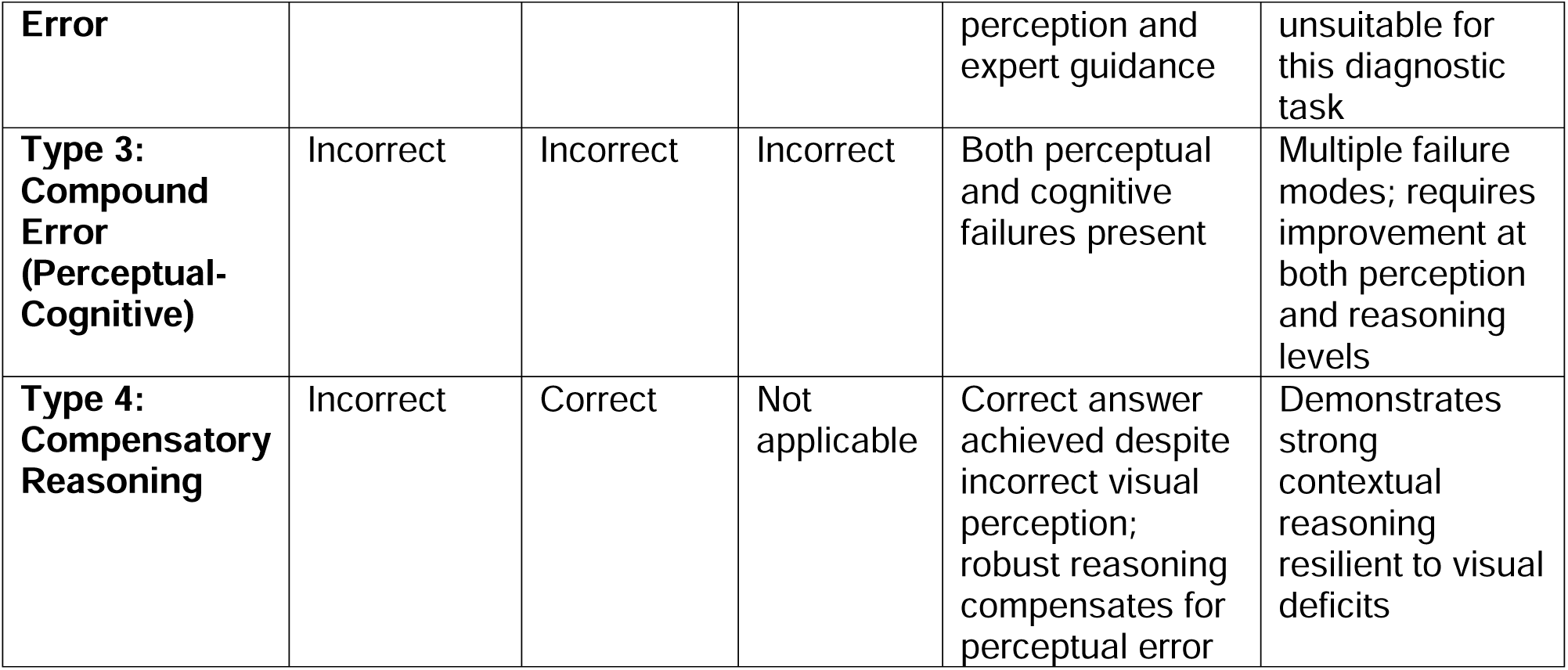
Error type classification.

A perceptual failure (Type 1) was recorded when a model produced both an inaccurate image description and an incorrect answer in Phase 1 but answered correctly once expert visual description was provided in Phase 2, indicating that the initial failure originated at the level of visual encoding, and that reasoning capacity was intact once accurate visual input was supplied.

A correctable cognitive error (Type 2A) was recorded when the model described the image accurately but answered incorrectly in Phase 1, yet corrected its response after expert guidance in Phase 2, suggesting a reasoning failure that is responsive to structured visual emphasis; a persistent cognitive error (Type 2B) was recorded when accurate perception in Phase 1 still produced an incorrect answer that remained incorrect even after Phase 2 intervention, indicating a fundamental reasoning deficit that expert visual input alone could not overcome.

A compound error (Type 3) reflected simultaneous perceptual and cognitive failure, with incorrect descriptions and answers persisting across both phases; and compensatory reasoning (Type 4) captured instances where correct answers were reached in Phase 1 despite inaccurate image descriptions, reflecting robust contextual reasoning resilient to visual deficits.

## STATISTICAL ANALYSIS

All statistical analyses were performed using SPSS Statistics version 21 (IBM Corporation, Armonk, NY, USA) and Python version [3.11] (Python Software Foundation). Statistical significance was set at α = 0.05 for all tests.

Baseline diagnostic accuracy and visual description accuracy were compared across LLMs using chi-square tests of independence. Linear regression assessed the effects of LLM model, image modality, and their interaction on diagnostic accuracy.

Intervention-induced improvement was assessed using McNemar’s test for paired binary data, comparing Phase 1 and Phase 2 accuracy within each LLM.

Error type distributions across LLMs were compared using chi-square test, with perceptual versus cognitive failure patterns examined by collapsing error types into perceptual v/s cognitive dominant types.

## RESULTS

### 1. Visual Description Accuracy

Visual perception performance varied significantly across models, χ² (5, N = 300) = 14.16, p = .015, indicating substantial architectural differences in multimodal image interpretation capabilities. Google Gemini 2.5 demonstrated superior visual description accuracy (41 correct, 82.0%), followed by GPT 5.0 (39 correct, 78.0%). GLM-4.6 achieved moderate visual accuracy (32 correct, 64.0%), while Claude Sonnet 4.5 and Perplexity Sonar both achieved 62.0% accuracy (31 correct each). Grok exhibited the lowest visual description performance (26 correct, 52.0%). (Table 2)

**Table 2.**
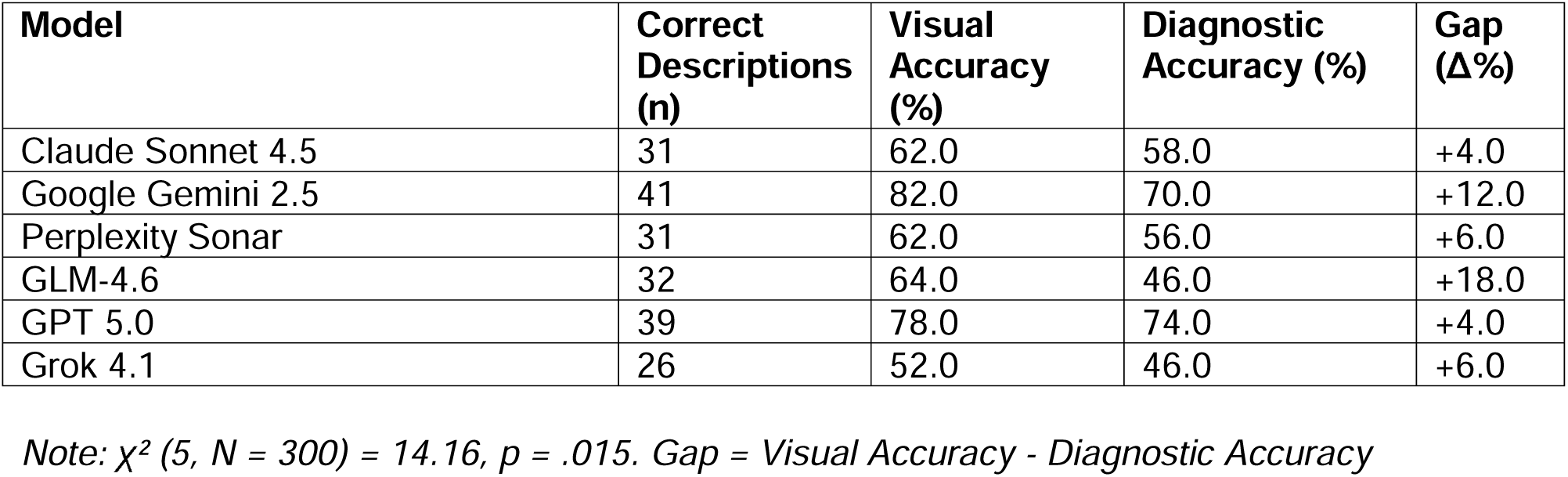
Visual Description Accuracy by LLM.

| Model | Correct Descriptions (n) | Visual Accuracy (%) | Diagnostic Accuracy (%) | Gap (Δ%) |
| --- | --- | --- | --- | --- |
| Claude Sonnet 4.5 | 31 | 62.0 | 58.0 | +4.0 |
| Google Gemini 2.5 | 41 | 82.0 | 70.0 | +12.0 |
| Perplexity Sonar | 31 | 62.0 | 56.0 | +6.0 |
| GLM-4.6 | 32 | 64.0 | 46.0 | +18.0 |
| GPT 5.0 | 39 | 78.0 | 74.0 | +4.0 |
| Grok 4.1 | 26 | 52.0 | 46.0 | +6.0 |

*Note: $\chi^2$ (5, N = 300) = 14.16, p = .015. Gap = Visual Accuracy - Diagnostic Accuracy*

### 2. PERFORMANCE

#### 2.1 Overall Performance

Six multimodal large language models completed 300 total evaluations across 50 AAP In-Service Examination questions. At baseline (first attempt), overall diagnostic accuracy ranged from 46.0% to 74.0% across models, spanning a 28-percentage-point range. GPT 5.0 achieved the highest baseline performance (37 correct responses, 74.0%), followed by Google Gemini 2.5 (35 correct, 70.0%). Claude Sonnet 4.5 and Perplexity Sonar demonstrated intermediate performance (29 and 28 correct, 58.0% and 56.0% respectively), while GLM-4.6 and Grok 4.1 exhibited the lowest baseline accuracy (23 correct each, 46.0%).(Table 3, Fig 3)

**Figure 3.**
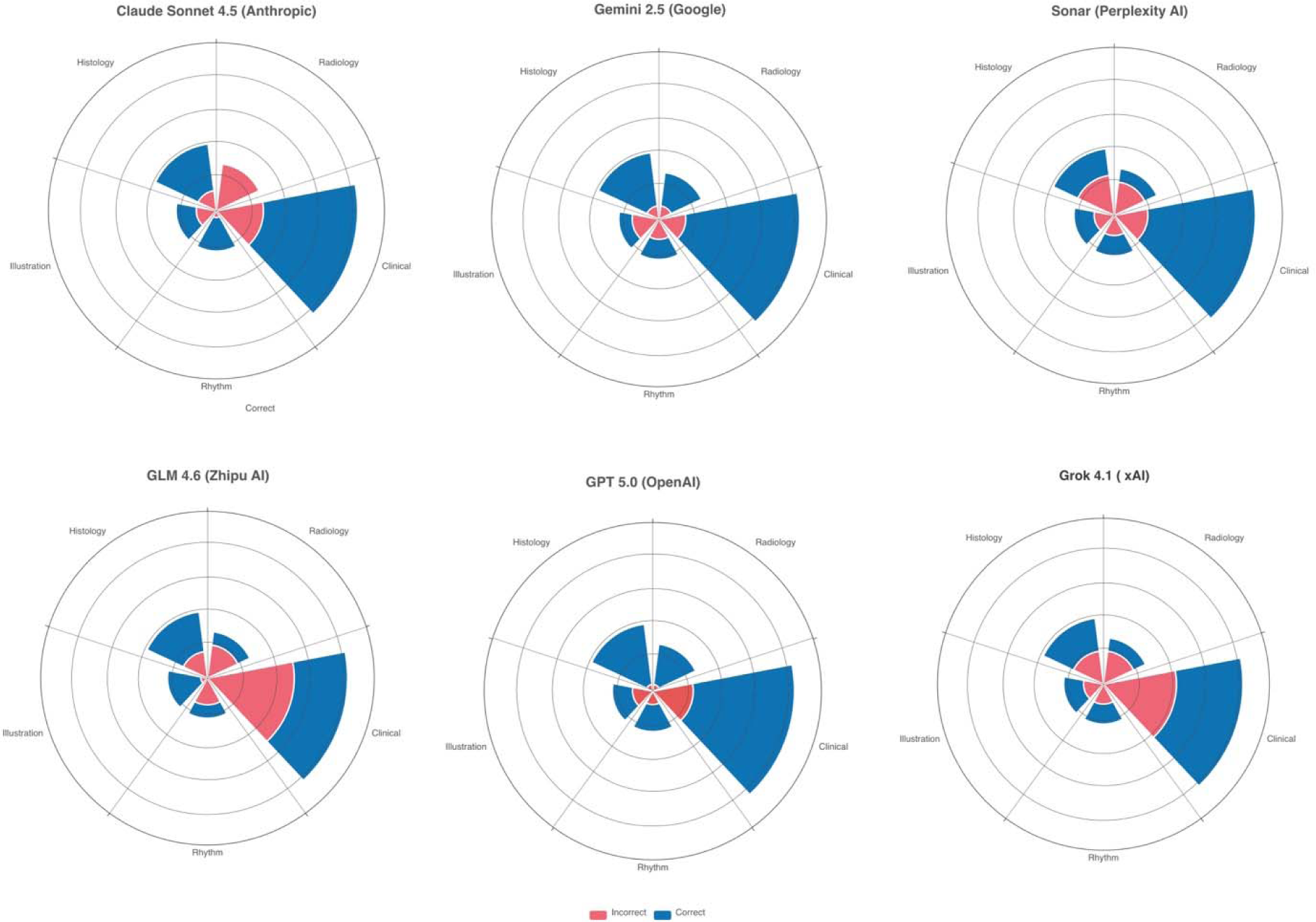
Baseline Diagnostic Accuracy by LLM and Image Domain. Nightingale rose diagrams showing Phase 1 performance distribution across five imaging modalities (Clinical photograph, Histopathology, Anatomical illustration, Radiology, Cardiac rhythm strip) for each LLM. Blue segments: correct responses; pink segments: incorrect responses. Segment area proportional to question count per domain.

**Table 3.** Baseline Diagnostic Accuracy by LLM (First Attempt).

| Model | Correct (n) | Incorrect (n) | Accuracy (%) |
| --- | --- | --- | --- |
| Claude Sonnet 4.5 | 29 | 21 | 58.0 |
| Google Gemini 2.5 | 35 | 15 | 70.0 |
| Perplexity Sonar | 28 | 22 | 56.0 |
| GLM-4.6 | 23 | 27 | 46.0 |
| GPT 5.0 | 37 | 13 | 74.0 |
| Grok 4.1 | 23 | 27 | 46.0 |

A chi-square test of independence revealed statistically significant differences in baseline accuracy among the six LLMs, χ² (5, N = 300) = 14.22, p = .014. This indicates that model architecture selection significantly influenced diagnostic performance at baseline, with top-performing models demonstrating 28 percentage points higher accuracy than the lowest performers.

#### 2.2 Linear Regression Analysis: LLM × Domain Interactions

Logistic regression was used to model item-level accuracy (correct = 1, incorrect = 0) as a function of LLM identity and question domain, including their interaction, to evaluate whether specific models behaved differently across imaging modalities. Claude Sonnet 4.5 and Histopathology were specified as reference categories, so all parameter estimates are interpreted relative to Claude Sonnet 4.5 answering Histopathology questions.

##### 2.2.1 Main Effects and Interaction

Global Score tests demonstrated that the LLM factor was statistically significant, χ² (5) = 14.22, p = .014, indicating that at least one model differed from Claude Sonnet 4.5 in overall odds of providing a correct answer across domains. The domain factor showed a trend toward significance, χ² (4) = 8.56, p = .073, suggesting modest differences in difficulty between imaging modalities once model effects were accounted for. Critically, the LLM × Domain interaction block was statistically significant, χ² (20) = 34.50, p = .023, confirming that relative performance of the LLMs varied by domain rather than following strictly parallel patterns across all question types.

##### 2.2.2 Model-Specific Performance

In all models, Claude Sonnet 4.5 was specified as the reference LLM and Histopathology as the reference domain, so all score tests are interpreted relative to Claude Sonnet 4.5 answering Histopathology questions.

Using Claude Sonnet 4.5 as the reference, global score tests indicated that at least one LLM differed from Claude Sonnet 4.5 in overall odds of a correct response (LLM factor, χ² (5) = 14.22, p = .014). The individual score test for the level coded as LLM 5 (GPT 5.0) was statistically significant (Score χ² (1) = 6.06, p = 0.014), indicating that GPT 5.0’s overall odds of a correct response differed from those of Claude Sonnet 4.5 after accounting for domain and their interaction. Gemini 2.5 (Score χ² (1) = 3.36, p = 0.067) and GLM-4.6 (Score χ² (1) = 3.76, p = 0.053) showed borderline evidence of differing from Claude Sonnet 4.5, whereas Perplexity Sonar did not differ significantly from Claude Sonnet 4.5 (Score χ² (1) = 0.13, p = 0.714), suggesting broadly comparable overall odds of correctness in the multivariable model.

Domain main effects (Anatomical illustration, Cardiac rhythm strip, Clinical photograph, Radiology versus Histopathology) all had p values greater than 0.05, indicating that no single domain was uniformly more or less difficult than Histopathology after adjusting for LLM differences (all individual domain score tests p ≥ .12). However, the significant LLM × Domain interaction (χ² (20) = 34.50, p = 0.023) underscores that the apparent difficulty of a given imaging domain depends on which LLM is responding.

##### 2.2.3 Domain-Specific Model Strengths and Weaknesses

The global LLM × Domain interaction was statistically significant (χ² (20) = 34.50, p = 0.023), indicating that relative model performance varied across imaging domains rather than following parallel patterns. Inspection of the individual interaction score tests showed that most LLM– domain combinations did not deviate significantly from what would be expected based on their main effects (all p ≥ 0.051), but two pairs stood out. (Table 4)

**Table 4.** Baseline Diagnostic Accuracy by LLM × Domain (%).

| Domain | GPT 5.0 | Claude Sonnet 4.5 | GLM -4.6 | Google Gemini 2.5 | Grok 4.1 | Perplexity Sonar |
| --- | --- | --- | --- | --- | --- | --- |
| Clinical | 71.4 | 66.7 | 38.1 | 81.0 | 47.6 | 76.2 |
| Histopathology | 90.0 | 70.0 | 60.0 | 80.0 | 50.0 | 40.0 |
| Anatomical illustration | 50.0 | 50.0 | 83.3 | 33.3 | 50.0 | 50.0 |
| Radiology | 85.7 | 0.0 | 28.6 | 71.4 | 28.6 | 28.6 |
| Cardiac rhythm strip | 66.7 | 83.3 | 33.3 | 50.0 | 50.0 | 50.0 |

**Note:** Values represent percentage of questions answered correctly at baseline (first attempt). Reference categories: Claude Sonnet 4.5 (LLM), Histopathology (domain). LLM main effect: $\chi^2 (5) = 14.22$ , $p = .014$ . Domain main effect: $\chi^2 (4) = 8.56$ , $p = .073$ . LLM × Domain interaction: $\chi^2 (20) = 34.50$ , $p = .023$ . Significant interactions: Gemini 2.5 × Clinical photograph ( $p = .029$ , $OR < 1$ ); GPT 5.0 × Histopathology ( $p = .039$ , $OR > 1$ ).

First, the interaction between Gemini 2.5 and the Clinical photograph domain (LLM 2 × Domain 4) was significant (Score χ² (1) = 4.75, p = 0.029), indicating that Gemini 2.5’s performance in Clinical imaging differed from what would be predicted from Gemini 2.5’s overall effect and the overall Clinical effect alone. In the observed accuracy table, Gemini 2.5 reached 81.0% correct in Clinical photograph but only 33.3% in Anatomical illustration, suggesting a profile that is comparatively stronger on Clinical photograph than on schematics.

Second, the interaction between GPT 5.0 and Histopathology (LLM 5 × Domain 1) was significant (Score χ² (1) = 4.27, p = 0.039), indicating that GPT 5.0’s performance in Histopathology likewise departed from the pattern implied by its main effect and the domain main effect. Consistent with this, GPT 5.0 achieved 90.0% accuracy on Histopathology questions, exceeding both Claude Sonnet 4.5’s 70.0% and Gemini 2.5’s 80.0% in that domain. All other LLM × Domain interactions had non-significant score tests (p ≥ .051), suggesting that domain-specific deviations from the average pattern were concentrated in these Gemini 2.5– Clinical photograph and GPT 5.0–Histopathology combinations.

Radiology exhibited the widest performance spread, ranging from 0.0% accuracy for Claude Sonnet 4.5 to 85.7% for GPT 5.0, an 85.7-percentage-point difference. By contrast, Anatomical illustration items showed a 50-point range, from 33.3% for Google Gemini 2.5 to 83.3% for GLM-4.6, with GLM-4.6 performing strongly on Anatomical illustrations (83.3%) but poorly on clinical photographs (38.1%). Claude Sonnet 4.5 demonstrated the opposite pattern, with excellent baseline performance on Cardiac rhythm strips (83.3%) but complete failure on radiology (0.0%).

These crossed performance profiles support the presence of a meaningful LLM × Domain interaction (χ² (20) = 34.50, p = 0.023) and emphasize that model choice should be tailored to the predominant imaging modality rather than inferred from a single aggregate accuracy metric.

### 3. Intervention-Induced Improvement Analysis

Expert visual description provision resulted in substantial and statistically significant improvements in diagnostic accuracy across all models and domains. Figure 4 illustrates the magnitude and distribution of improvement by LLM and domain.

**Figure 4.**
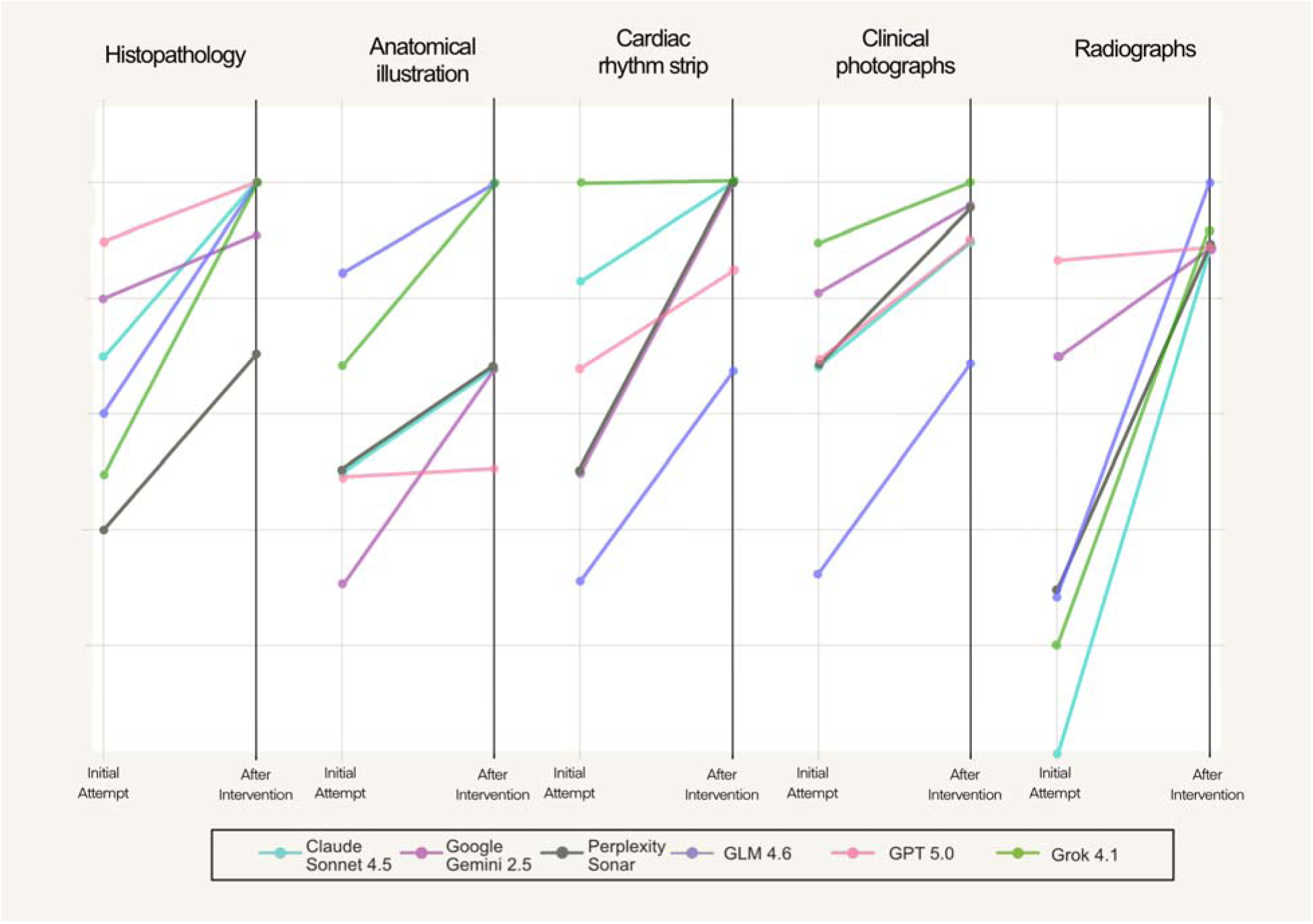
Domain-Specific Improvement Following Expert Visual Correction. Slope graphs showing baseline (Initial Attempt) and post-intervention (After Intervention) diagnostic accuracy for each of the five imaging domains (Histopathology, Anatomical illustration, Cardiac rhythm strip, Clinical photographs, Radiology). Each colored line represents one LLM’s performance trajectory within that domain, connecting baseline accuracy to post-intervention accuracy. Y-axis represents percentage correct; X-axis shows assessment phase (Initial Attempt vs. After Intervention).

**Figure 4a.**
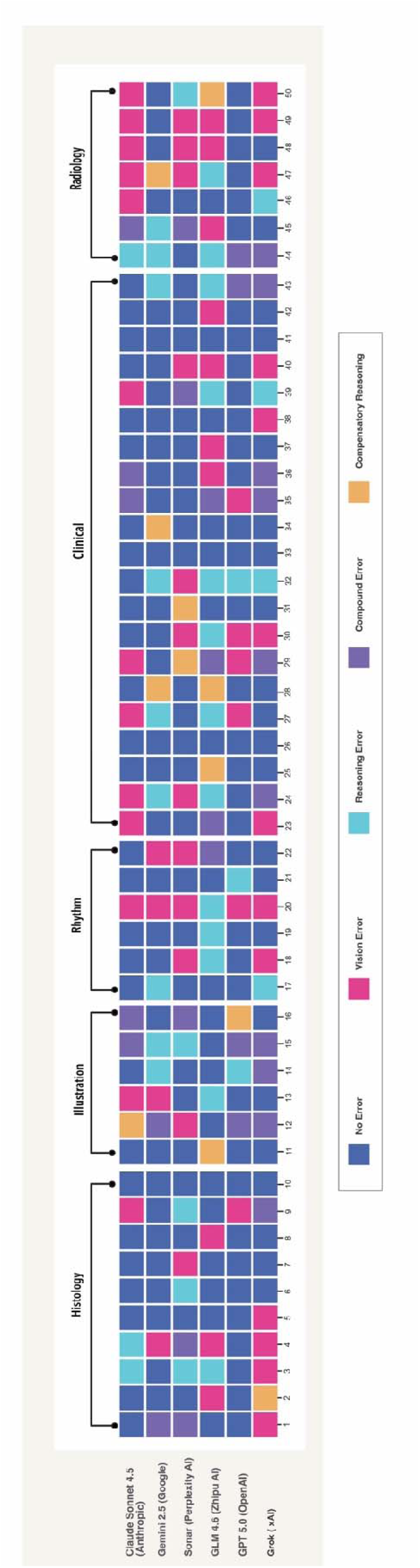
Error Type Distribution by LLM and Image Domain. Heatmap showing error classification patterns across all 50 questions (columns 1-50) grouped by imaging domain (Histopathology, Anatomical illustration, Cardiac rhythm strip, Clinical photographs, Radiology) for each of the six LLMs (rows). Error types: No Error, Vision Error (perceptual failure), Reasoning Error (cognitive failure), Compound Error (both perceptual and cognitive failures), and Compensatory Reasoning (correct answer despite incorrect visual description). Each cell represents one question-model evaluation classified using Norman’s dual-process error taxonomy.

#### 3.1 Domain-Specific Improvement

Domain type significantly influenced baseline diagnostic difficulty and post-intervention performance. At baseline, domain-level accuracy varied significantly, χ² (4, N = 300) = 14.69, p = 0.005, with Radiology demonstrating the lowest accuracy (40.0%, 24 of 60 correct) and clinical photographs the highest (71.7%, 43 of 60 correct). Following intervention, domain differences persisted, χ² (4, N = 300) = 11.78, p = 0.019, though overall accuracy increased substantially across all modalities.

Radiology questions showed the greatest absolute improvement, increasing from 40.0% to 91.7% (+51.7 percentage points), with 31 of 36 initially incorrect responses corrected. Clinical photographs showed the smallest improvement (+20.0 points, from 71.7% to 91.7%), consistent with a ceiling effect given high baseline performance. Anatomical illustration items improved from 55.0% to 76.7% (+21.7 points) but remained the lowest performing domain post-intervention, with 14 of 60 attempts still incorrect despite expert visual descriptions. (Table 5, Figure 4)

**Table 5.** Domain-Level Accuracy Before and After Intervention.

| Domain | Baseline Correct (n) | Baseline (%) | Post-Intervention Correct (n) | Post-Intervention (%) | Improvement ($\Delta\%$) |
| --- | --- | --- | --- | --- | --- |
| Clinical photograph | 43 | 71.7 | 55 | 91.7 | +20.0 |
| Histopathology | 39 | 65.0 | 56 | 93.3 | +28.3 |
| Anatomical illustration | 33 | 55.0 | 46 | 76.7 | +21.7 |
| Radiology | 24 | 40.0 | 55 | 91.7 | +51.7 |
| Cardiac rhythm strip | 38 | 63.3 | 55 | 91.7 | +28.4 |

*Note: N = 60 per domain (6 LLMs × domain-specific question count). Baseline: $\chi^2 (4) = 14.69$ , $p = .005$ . Post-intervention: $\chi^2 (4) = 11.78$ , $p = .019$ .*

#### 3.2 Model-Specific Improvement

Paired pre-post-performance was evaluated using McNemar’s test on per-item correctness for each LLM. Across all models, there was a statistically significant increase in the proportion of correct responses from baseline to post-intervention (all p < .05). Among items that were initially incorrect, the proportion corrected after expert visual description ranged from 46.2% for GPT 5.0 (6 of 13 incorrect items) to 76.2% for Claude Sonnet 4.5 (16 of 21 incorrect items), indicating that the intervention consistently improved item-level accuracy with varying magnitudes of benefit across models.(Table 6, Fig 5)

**Figure 5.**
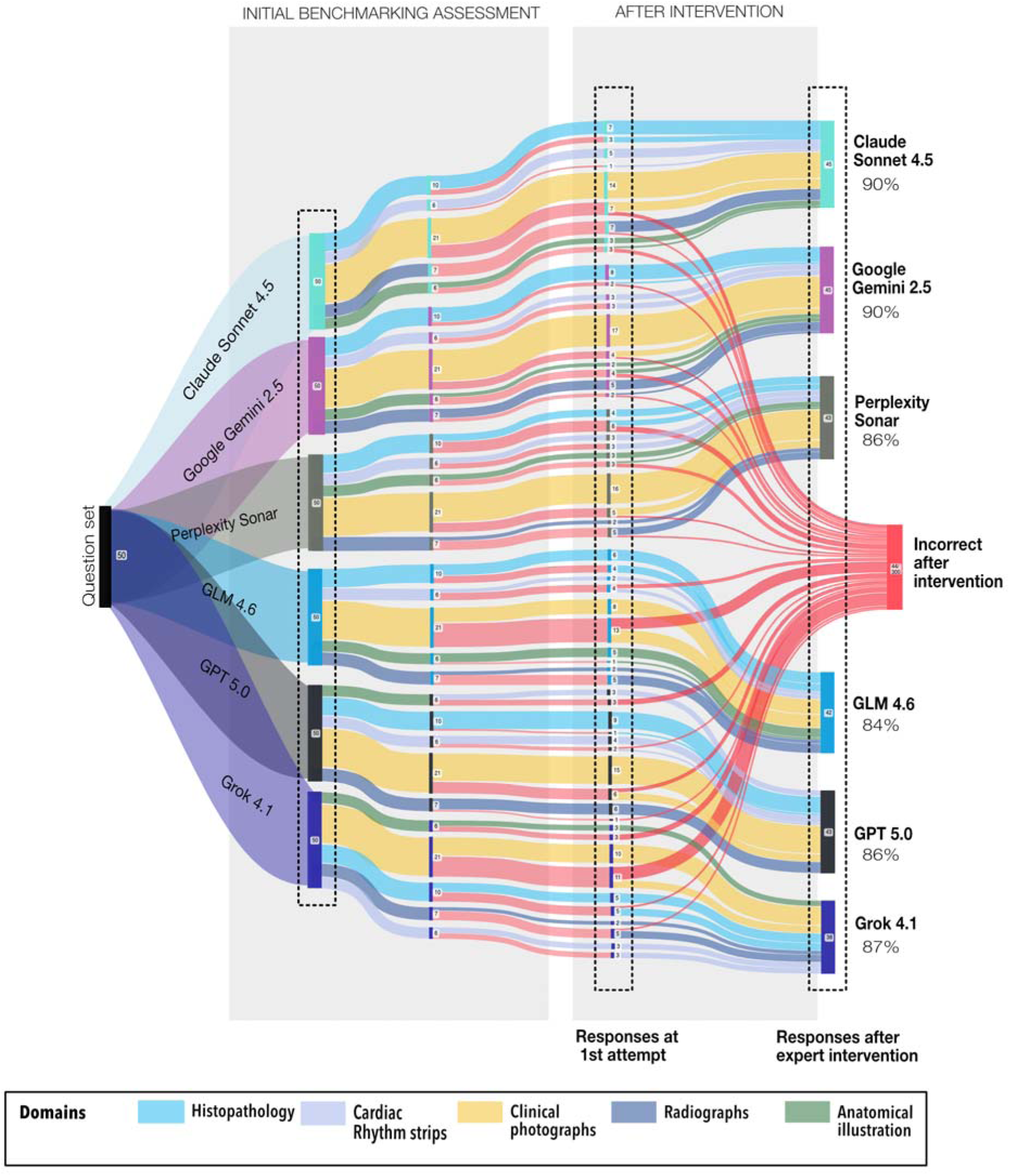
Diagnostic Performance Flow Across Initial Assessment and Expert-Guided Intervention. Sankey diagram illustrating response trajectories for all six LLMs across 50 questions (300 total evaluations). Left panel shows initial benchmarking assessment performance by LLM (question distribution pie chart) and baseline accuracy. Middle columns display Phase 1 responses color-coded by imaging domain (Histopathology: light blue; Cardiac rhythm strip: purple; Clinical photographs: yellow; Radiology: dark blue; Anatomical illustration: green). Right panel shows Phase 2 outcomes following expert visual correction, with flows indicating responses that remained correct, became correct after intervention, or remained incorrect. Red flows on far right represent persistent errors despite expert-guided correction.

**Table 6.** Model-Specific Improvement Following Intervention.

| **LLM** | **Baseline Correct** | **Post-Intervention Correct** | **Items Corrected (n)** | **Correction Rate (%)** | **McNemar $\chi^2$** | **p-value** |
| --- | --- | --- | --- | --- | --- | --- |
| Claude Sonnet 4.5 | 29 | 45 | 16/21 | 76.2 | 14.06 | <.001 |
| Google Gemini 2.5 | 35 | 45 | 10/15 | 66.7 | 8.10 | .004 |
| Perplexity Sonar | 28 | 43 | 15/22 | 68.2 | 13.07 | <.001 |
| GLM-4.6 | 23 | 42 | 19/27 | 70.4 | 17.05 | <.001 |
| GPT 5.0 | 37 | 43 | 6/13 | 46.2 | 4.17 | .041 |
| Grok 4.1 | 23 | 38 | 15/27 | 55.6 | 13.07 | <.001 |

*Note: Correction rate = proportion of initially incorrect items that became correct after intervention. All models showed significant improvement ( $p < .05$ ).*

### 4. Error Classification and Mechanistic Failure Patterns

To examine whether different LLMs exhibited qualitatively distinct failure modes beyond overall accuracy differences, each response was classified using an extended dual-process error taxonomy based on Norman’s model. This framework distinguished perceptual failures (vision-only errors), cognitive failures (reasoning-only errors), combined failures (compound errors), and compensatory successes (correct answers despite incorrect visual descriptions).(Table 1)

#### 4.1 Global Error Profile Analysis

An initial chi-square test of independence examined whether the full five-category error distribution differed across the six LLMs. The 6 (LLM) × 5 (error type) contingency table yielded χ² (20) = 29.9, p = .07, indicating only weak evidence that the detailed error taxonomy varied systematically by model at conventional significance thresholds. This suggested that fine-grained error distinctions (separating reasoning-only from compound errors, for example) did not reveal clear model-specific patterns with the current sample size. (Table 7, Fig 4)

**Table 7.**
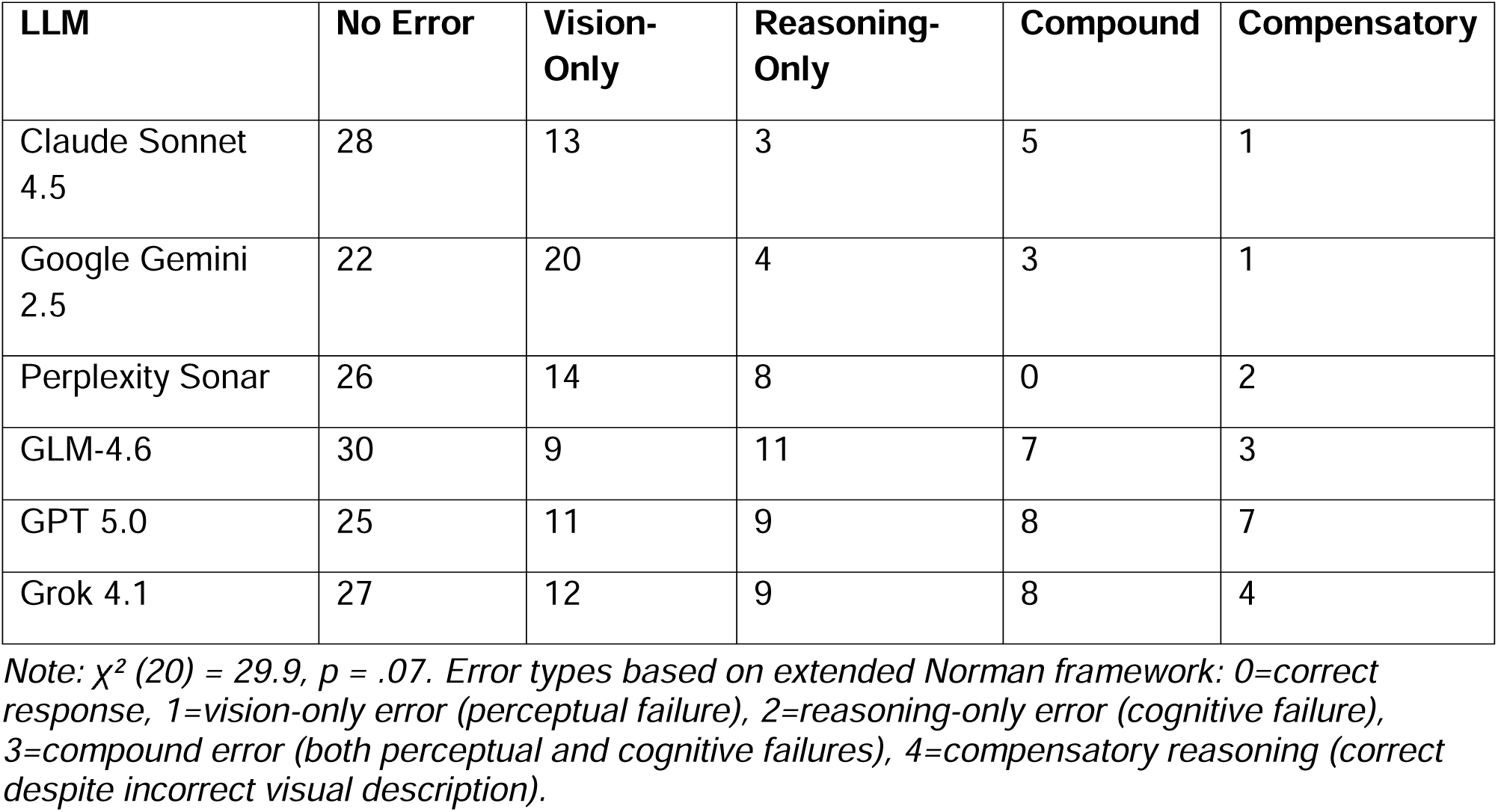
Error Type Distribution by LLM.

To explore whether different large language models exhibited distinct failure modes, we compared the distribution of error types across models using a chi-square test of independence. The contingency table crossed LLM (Claude Sonnet 4.5, Gemini, Sonar, GLM-4.6, GPT 5.0, Grok 4.1) with error classification (no error, vision-only error, reasoning-only error, compound error, compensatory reasoning). The Pearson chi-square statistic was χ² (20) ≈ 29.9, p ≈ 0.07. Thus, we did not find statistically significant evidence that the pattern of error types differed across models at the conventional α = 0.05 level. Although some models appeared to show qualitatively different mixes of vision, reasoning, compound, and compensatory errors, these differences did not reach statistical significance in this sample, suggesting that, within current power constraints, the models do not demonstrate clearly distinct error-pattern profiles (perceptual vs. cognitive) beyond chance variation.

This analysis was motivated by the hypothesis that some models might fail primarily at the perceptual (vision) level, whereas others might show predominantly cognitive (reasoning) or mixed perceptual–cognitive failures. The chi-square test, however, did not support strong model-specific error profiles, implying that, given the present dataset, the relative balance of perceptual, reasoning, compound, and compensatory errors is broadly similar across LLMs.

#### 4.2 Focused Analysis: Perceptual vs. Cognitive Failure Modes

To directly address the mechanistic question of whether some LLMs fail primarily at visual perception while others fail at downstream reasoning, the error taxonomy was collapsed to a binary distinction among incorrect responses only: perceptual errors (vision-only) versus all other error types (reasoning-only, compound, compensatory).

A Pearson chi-square test revealed a significant association between model and error mechanism, χ² (5) = 17.60, p = .003, Cramer’s V = 0.320. Examination of standardized residuals showed that Gemini exhibited the strongest perceptual bias (standardized residual +2.0), with 71.4% of its errors classified as vision-only (20 of 28 errors), consistent with a predominantly perceptual failure mode. Conversely, GPT-5.0 and GLM-4.6 demonstrated the opposite pattern (standardized residuals −1.3 and −1.3), with approximately 70% of their errors classified as non-perceptual (ChatGPT: 24 of 35 errors; GLM-4.6: 21 of 30 errors), suggesting these models’ failure loci lie primarily in cognitive or integrative processing. Claude Sonnet, Perplexity Sonar, and Grok exhibited more balanced error profiles with perceptual errors comprising 36-59% of failures and standardized residuals near zero.

## DISCUSSION

This study benchmarked six multimodal large language models on image-based periodontal examination questions, revealing substantial variation in diagnostic accuracy, differential responsiveness to expert visual correction, and qualitatively distinct failure mechanisms across models. Collectively, the findings suggest that current multimodal LLM performance in this domain is shaped not only by overall architectural capability, but by the interplay between visual encoding fidelity, domain-specific reasoning, and the nature of the imaging modality presented. This pattern mirrors the findings of Lau et al., who demonstrated in the VQA-RAD dataset that clinically generated questions vary systematically in difficulty across anatomical regions and imaging modalities, underscoring the inadequacy of single-metric benchmarking in medical visual question answering. (20)

Our present findings demonstrate that multimodal LLM diagnostic performance in periodontal image interpretation is both substantial in its variability and highly dependent on imaging context, challenging the premise that a single model can serve as a universal tool for clinical education. Similar model-based and context-specific variability has been reported across other medical imaging domains, where model performance rankings consistently shift depending on imaging modality, anatomical region, and clinical task.(1,21)

At baseline, accuracy ranged from 46.0% to 74.0% across the six models. This 28-percentage-point gap is consistent with prior benchmarking studies in medical imaging that have documented comparable inter-model variability across radiology, pathology, and ophthalmology tasks. Critically, however, aggregate accuracy alone proved misleading: the significant LLM × domain interaction (χ² (20) = 34.50, *p* = 0.023) confirmed that model rankings were not stable across imaging contexts, a pattern also observed in prior medical VQA benchmarking where question difficulty varied systematically by imaging modality and anatomical region. (22)

Specific interaction terms revealed non-uniform performance patterns. The Gemini 2.5 × Clinical photographs interaction had a significant negative coefficient (p = .029, OR < 1), indicating that Gemini’s odds of a correct response in the Clinical photograph domain were lower than expected based on its overall performance, consistent with a domain-specific weakness. Conversely, the GPT 5.0 × Histopathology interaction showed a significant positive coefficient (p = .039, OR > 1), implying that GPT 5.0 performed relatively better in Histopathology than its overall main effect would predict, partially compensating for its overall disadvantage relative to Claude Sonnet 4.5. This finding aligns with Panagoulias et al., who reported approximately 84% accuracy for GPT-4V on pathology image-based multiple-choice questions, suggesting that GPT-family models may carry a consistent advantage in histopathological reasoning. (23) Meanwhile, GLM-4.6, a comparatively lower overall performer, led all models in Anatomical illustration-based items (83.3%), suggesting that architectural differences in spatial reasoning or training data composition may confer domain-specific advantages that aggregate benchmarks obscure. Importantly, no individual domain main effect reached significance (all p ≥ .12), confirming that apparent domain difficulty is not a fixed property but rather an LLM-dependent one.

Overall, these crossed performance profiles are consistent with Zhang et al.’s finding in the PMC-VQA benchmark that models pre-trained on diverse medical imaging datasets show markedly different strengths across modality types, suggesting that training data composition and vision encoder design shape domain-specific competencies in ways that overall accuracy metrics fail to capture. (22) Taken together, these findings support a domain-stratified strategy for LLM selection in dental and medical education: rather than relying solely on models with the highest overall accuracy, educators and clinicians should prioritize models whose performance is optimized for the imaging modality that predominates in their specific clinical or educational context, a principle that is consistent with prior work on modality-specific and domain-adapted AI systems.(17,24,25)

A central finding of this study is that expert-guided visual correction produced statistically significant improvements in diagnostic accuracy across all six models, with McNemar tests confirming pre-to-post intervention gains in every case (all *p* < .05). This result establishes that real-time provision of accurate visual descriptions can function as an effective corrective mechanism in multimodal LLM diagnostic workflows, a principle conceptually analogous to the Woodpecker hallucination correction framework proposed by Yin et al., in which externally supplied visual knowledge was used to realign model outputs with image content achieving F1-score gains of up to 30.66 percentage points over MiniGPT-4 on the POPE object-level hallucination benchmark, without requiring model retraining. (26) However, the magnitude of improvement varied substantially across models in ways that are not reducible to baseline accuracy alone, and these differential correction rates carry architectural implications. Similar evidence comes from a recent evaluation of multimodal reasoning models in radiology, in which the integration of descriptive-text inputs substantially increased diagnostic accuracy across all models, with the best-performing model improving from 56.2% on imaging alone to 71.2% with structured text input, while model architecture remained the fundamental determinant of baseline performance, with prompting strategies modulating efficiency within those architectural constraints.(27)

Claude Sonnet 4.5 demonstrated the highest correction rate (76.2%; 16 of 21 initially incorrect items corrected) despite only moderate baseline accuracy (58.0%), indicating that its diagnostic reasoning is capable of capitalizing on clarified visual input and that initial failures predominantly reflected vision-encoding constraints rather than reasoning deficits. GPT-5.0, by contrast, showed the lowest correction rate (46.2%; 6 of 13 items corrected) despite the highest baseline accuracy (74.0%), suggesting that its residual errors after intervention represent reasoning or cross-modal integration failures that persist regardless of visual input quality. The visual description accuracy gap further illuminates these differences: GLM-4.6 exhibited an 18-percentage-point gap between visual accuracy (64.0%) and diagnostic accuracy (46.0%), suggesting substantial reasoning inefficiency even when perception was adequate, whereas GPT-5.0 and Claude Sonnet 4.5 showed gaps of only 4 percentage points each, indicating tighter coupling between perceptual and reasoning processes. Beyond aggregate accuracy differences, binary error classification revealed qualitatively distinct failure mechanisms across models, implicating different architectural loci of dysfunction with direct relevance to targeted model improvement. Applying the dual-process error taxonomy described in the Methods, errors were collapsed into perceptual-dominant and cognitive-dominant profiles to assess whether systematic inter-model differences existed in failure mechanism rather than failure rate alone. (18,19,28)

This distinction is architecturally meaningful: in multimodal LLMs, visual encoding and language-based reasoning operate through largely separable components, the vision encoder and multimodal connector on one hand, and the transformer-based language model on the other, such that failures at each stage carry different implications for model refinement. Model-level error distributions reflect this separability. Gemini 2.5 exhibited a predominantly perceptual failure profile, with 71.4% of its errors classified as vision-only (standardized residual = 2.0), consistent with systematic limitations in visual feature extraction despite preserved downstream reasoning capacity, evidenced by its high correction rate following expert visual description.

GPT-5.0 and GLM-4.6 demonstrated inverse patterns, with approximately 69% and 70% of their errors respectively classified as non-perceptual (standardized residuals = −1.3 each), indicating that failure in these models resides primarily in reasoning or cross-modal integration processes. Domain-level trajectories corroborate this interpretation: the dramatic Radiology improvement (+51.7 percentage points) reflects a predominantly perceptual bottleneck in that domain, while Anatomical illustration-based items remained the lowest-performing post-intervention (76.7%), suggesting reasoning demands involving spatial inference and schematic interpretation that visual realignment alone cannot resolve. Collectively, these patterns suggest that correction responsiveness is itself an architecturally determined property, distinct from baseline accuracy, and one that aggregate benchmarking alone would fail to differentiate. This is consistent with evidence that vision encoder performance on general-domain benchmarks does not reliably predict VLM diagnostic capability, and that task-specific encoder adaptation yields performance gains independent of backbone scale (29). Architectural investment in vision encoder fidelity may therefore yield the greatest gains for perceptual-dominant models, while cognitive-dominant failure profiles may be better addressed through structured reasoning decomposition or domain-specific fine-tuning.(30–32)

Rather than deploying these systems autonomously, an expert-in-the-loop model, one in which clinician-provided structured visual descriptions guide model reasoning in real time may substantially outperform fully autonomous deployment, particularly for models with strong reasoning capacity but weak visual encoding. This framework aligns with broader calls in the medical AI literature for interactive rather than autonomous AI in high-stakes diagnostic contexts, and points toward the practical utility of structured visual reporting templates that systematically surface image findings before LLM reasoning is engaged.(33–35) The divergent failure profiles documented here raise the broader possibility that ensemble approaches routing questions to domain-optimized models may outperform single-model deployment, a hypothesis that warrants prospective evaluation across larger and more diverse imaging datasets and live clinical workflows.(1,36)

Together, these patterns offer a tentative but internally consistent picture: the mechanisms underlying multimodal diagnostic failure may differ substantially across architectures and are likely not fully captured by overall accuracy alone. Correction rate provides a complementary perspective that could help differentiate models with similar aggregate performance but distinct error profiles, suggesting that clinical deployment decisions may benefit from considering failure mode distribution alongside headline accuracy metrics, an avenue that warrants further systematic investigation.

### Clinical Implementation Considerations

The performance profiles documented in this study, considered alongside the demonstrated responsiveness of multimodal LLMs to expert visual guidance, suggest several practical applications in dental and medical education. Multimodal LLMs accessible through consumer interfaces represent a readily available resource for trainees seeking on-demand image interpretation feedback outside supervised clinical settings, and the present findings suggest their utility is maximized when approached as interactive reasoning partners rather than authoritative references, with discrepancies between model outputs and established answers used as active learning prompts. The substantial improvement in diagnostic accuracy following expert visual description provision supports the development of structured educational frameworks in which validated visual description templates are paired with LLM reasoning prompts, guiding trainees and students through systematic image observation before answer selection, an approach that aligns with established pedagogical principles of structured visual analysis in radiology and pathology education, and may be particularly valuable in settings where real-time expert supervision is limited. The domain-specific difficulty gradients observed across models, with Radiology and Anatomical illustration-based items proving most resistant to correct interpretation, converge with established patterns of imaging domain difficulty in standardized assessment, suggesting that systematic LLM benchmarking may offer a reproducible complement to conventional item analysis in examination development.

### Limitations

The present study employed 50 image-based questions systematically selected from AAP In-Service Examination materials spanning 2012 to 2025, providing a clinically grounded evaluation context with established educational validity within periodontal specialty training.

Domain-specific sample sizes nonetheless ranged from 6 to 21 questions per modality, and while sufficient to detect large effects and significant interactions, these counts preclude definitive conclusions regarding small effect sizes in individual LLM-domain combinations. The findings are specific to periodontal and dental imaging contexts, and extension to other medical specialties warrants independent investigation. Models were accessed through consumer-facing interfaces by design, reflecting the real-world conditions under which students and residents engage with these tools and ensuring ecological validity for the educational context studied.

This approach necessarily precludes control over model parameters, system configurations, and version updates, and results should be interpreted as representative of a specific access period rather than fixed architectural capabilities.

Expert visual descriptions were developed by a board-certified periodontist (P.D.) and reviewed and approved by a second independent board-certified periodontist (R.H.), providing a clinically grounded and expert-validated reference standard. Accuracy of LLM-generated visual descriptions was evaluated against a pre-established expert-validated reference standard, anchoring all judgments to an objective benchmark rather than real-time clinical impression.

Accuracy was operationalized as a binary outcome, correct or incorrect, which substantially reduces the interpretive variability inherent to graded or qualitative rating schemes, yielding lower expected disagreement rates than scaled or interpretive assessments. To assess intra-rater reliability, the primary evaluator independently re-rated all descriptions four weeks after the initial assessment, with items presented in randomized order to minimize recall bias. Cohen’s kappa indicated almost perfect agreement between rating occasions (κ > 0.80), supporting the consistency and reproducibility of the binary accuracy classifications. Future studies should consider incorporating dual-rater or multiple-rater protocols with formal inter-rater agreement statistics to further strengthen visual description accuracy frameworks in multimodal LLM benchmarking contexts.

Phase 2 prompts were administered within the same conversation thread as Phase 1 to preserve ecological validity, though this introduces potential context carryover effects. The five-category error taxonomy analysis was exploratory given current sample size constraints (χ² (20) = 29.9, *p* = 0.07), and the binary perceptual versus cognitive collapse used for primary mechanistic inference represents a pragmatic and theoretically grounded analytic decision rather than a fundamental limitation of the framework. Additionally, consumer-facing interfaces may incorporate implicit in-context learning across conversation turns, such that Phase 2 responses may reflect accumulated conversational context beyond the expert description alone.

Finally, the possibility that some examination questions were present in model pre-training corpora cannot be excluded, and performance differences across models may partially reflect differential exposure to examination content rather than genuine reasoning capability. A central objective of this study was not only to quantify multimodal LLM diagnostic accuracy, but to probe the mechanisms underlying failure. To this end, the expert visual description intervention was designed as a diagnostic tool: by isolating the contribution of visual encoding from downstream reasoning, differential improvement patterns across domains and models can be used to infer where diagnostic pipeline errors originate. While causal attribution from a single intervention remains inherently inferential, the consistency of patterns across six models and four image domains provides convergent evidence for distinguishing predominantly perceptual failures, those resolved by accurate image description from predominantly cognitive failures, which persist regardless of visual input quality.

The observed patterns suggest that some failures were more readily corrected after visual clarification, particularly in Radiology, whereas Anatomical illustration items remained relatively challenging even after intervention. At the model level, correction rates varied across models, but these differences should be interpreted as sample-specific observations rather than stable architectural properties.

### Conclusions

This study demonstrates that multimodal LLM performance in periodontal image interpretation varies substantially across models and Clinical photograph domains, with no single architecture achieving consistent diagnostic reliability. The significant accuracy gains following expert visual correction observed across all models indicate that current failures are predominantly perceptual in nature, reflecting limitations in visual feature extraction rather than deficits in underlying periodontal knowledge. Critically, models exhibit distinct failure mode profiles: some are more susceptible to perceptual misclassification while others demonstrate systematic cognitive reasoning errors, suggesting that architectural interventions must be model-specific rather than universal. These findings collectively underscore that responsible clinical deployment of multimodal AI in periodontology requires task-specific model selection, structured human oversight at the visual interpretation stage, and continuous prospective validation against evolving clinical standards. As human-AI collaborative diagnostic systems move closer to clinical integration, the development of domain-adapted models trained on verified periodontal imaging corpora represents a necessary next step. Beyond technical development, however, the safe incorporation of these tools into clinical and educational practice demands clear institutional frameworks, including policies on AI-assisted diagnosis, formal training in critical appraisal of LLM outputs, and regulatory guidance that reflects the unique risks of high-stakes visual decision-making in healthcare.

## Data Availability

All data generated in this study are included in the Supporting Information files, including the expert-generated diagnostic descriptions, the multimodal large language model outputs, and associated tabulated results.

## Notes

### Competing Interest Statement

The authors have declared no competing interest.

